# The Polygenic Score Catalog: new functionality and tools to enable FAIR research

**DOI:** 10.1101/2024.05.29.24307783

**Authors:** Samuel A. Lambert, Benjamin Wingfield, Joel T. Gibson, Laurent Gil, Santhi Ramachandran, Florent Yvon, Shirin Saverimuttu, Emily Tinsley, Elizabeth Lewis, Scott C. Ritchie, Jingqin Wu, Rodrigo Canovas, Aoife McMahon, Laura W. Harris, Helen Parkinson, Michael Inouye

**Author notes:** Correspondence to* &. Denotes equal contributions (Samuel A. Lambert, Benjamin Wingfield).

## Abstract

Polygenic scores (PGS) have transformed human genetic research and have multiple potential clinical applications, including risk stratification for disease prevention and prediction of treatment response. Here, we present a series of recent enhancements to the PGS Catalog (www.PGSCatalog.org), the largest findable, accessible, interoperable, and reusable (FAIR) repository of PGS. These include expansions in data content and ancestral diversity as well as the addition of new features. We further present the PGS Catalog Calculator (`pgsc_calc`, https://github.com/PGScatalog/pgsc_calc), an open-source, scalable and portable pipeline to reproducibly calculate PGS that securely democratizes equitable PGS applications by implementing genetic ancestry estimation and score normalization using reference data. With the PGS Catalog & calculator users can now quantify an individual’s genetic predisposition for hundreds of common diseases and clinically relevant traits. Taken together, these updates and tools facilitate the next generation of PGS, thus lowering barriers to the clinical studies necessary to identify where PGS may be integrated into clinical practice.

## Main Text

### Expanding the PGS Catalog data content and evolving the user interface

The emergence of large cohorts of individuals with detailed phenotypic and genotype information has enabled the mapping of genetic variants across the genome associations with many hundreds of diseases and quantitative traits in genome-wide association studies (GWAS).^1^ This GWAS information, either individual-level or as summary statistics, can be leveraged to estimate an individual’s genetic predisposition for a given phenotype, represented as a score (weighted sum of variant dosages multiplied by their effect sizes) and is referred to as a polygenic score (PGS), polygenic risk score (PRS) or genetic/genomic risk score (GRS) depending on the context and application. Many tools exist to develop PGS (*i*.*e*. selecting the variants and weights included in the score), among the most common are Pruning+Thresholding/PRSice^2^, LDpred(2)^3^, or PRS-CS(x)^4^, and pipelines exist to automate development (e.g. GenoPred^5^). PGS have multiple research uses (e.g. instruments to interrogate molecular pathways or in gene x environment studies^6^) and potential clinical applications (e.g. improving risk-stratification, predicting treatment response, disease subtyping).^7^

The PGS Catalog (www.pgscatalog.org) was developed to collect and distribute information about PGS that have been developed so that analyses can be reproduced and the scores reused.^8^ The PGS Catalog is an open and FAIR (Findable, Accessible, Interoperable, Reusable) database, cataloging the relevant metadata required for accurate application and evaluation in a standardized format that is endorsed by co-developed reporting standards.^8,9^ This includes information about the samples and methods of PGS development procedure, performance metrics summarizing the predictive ability of the PGS in external samples, and a scoring file that can be applied to new samples to calculate the PGS. The PGS Catalog is based on data curated from publications as well as data deposited by authors directly.

Metadata and scoring files in the PGS Catalog are accessible by a web interface, FTP or REST API for programmatic access and download. The PGS Catalog has been visited by ∼27,000 users from over 140 countries in the past year and it serves as a platform for PGS research, for example as the source for scores which can be ensembled for improved polygenic prediction ^10^ or for return of PGS results to individuals as part of a clinical trial.^11^

Since the initial publication of the PGS Catalog, the data content has grown substantially and, as of May 8, 2024, the Catalog contains 4,735 PGS (a 721% increase) comprising data from 618 publications (519% increase) (**Figure 1**). Eligible publications are systematically identified using a machine-learning based literature triage^12^, trained on 1,704 publications that have been screened for eligibility since 2019 (encompassing 896 eligible PGS publications from 2008 - present), and re-trained yearly. The literature triage identifies ∼16 PGS Catalog eligible papers per week (out of ∼30 for screening), papers are prioritized for curation and data extraction if the scoring file is made available and if they include ancestrally diverse data or new traits. Processes for submissions have been streamlined, and authors are encouraged to submit directly to the Catalog by sending a scoring file and spreadsheet with required metadata about score development and evaluation (see www.pgscatalog.org/submit). Authors are now also provided the opportunity to embargo data until publication of the final manuscript in order to add PGS Catalog identifiers to their text (to comply with journal data sharing requirements, such as *Nature & Cell* groups). Data from preprints continue to meet publication eligibility and scores with licenses that do not require managed access are indexed. Within the last year, 23% of publications had data submitted directly by authors, and we continue to work with journals to ensure all PGS publications make their data accessible at the time of publication. The scores in the PGS Catalog predict traits mapping to 654 unique ontology terms^13^ (419% increase), with many recent traits being for less prevalent diseases (e.g. hypothyroidism) or quantitative traits (e.g. neuroimaging-derived traits).

**Figure 1.**
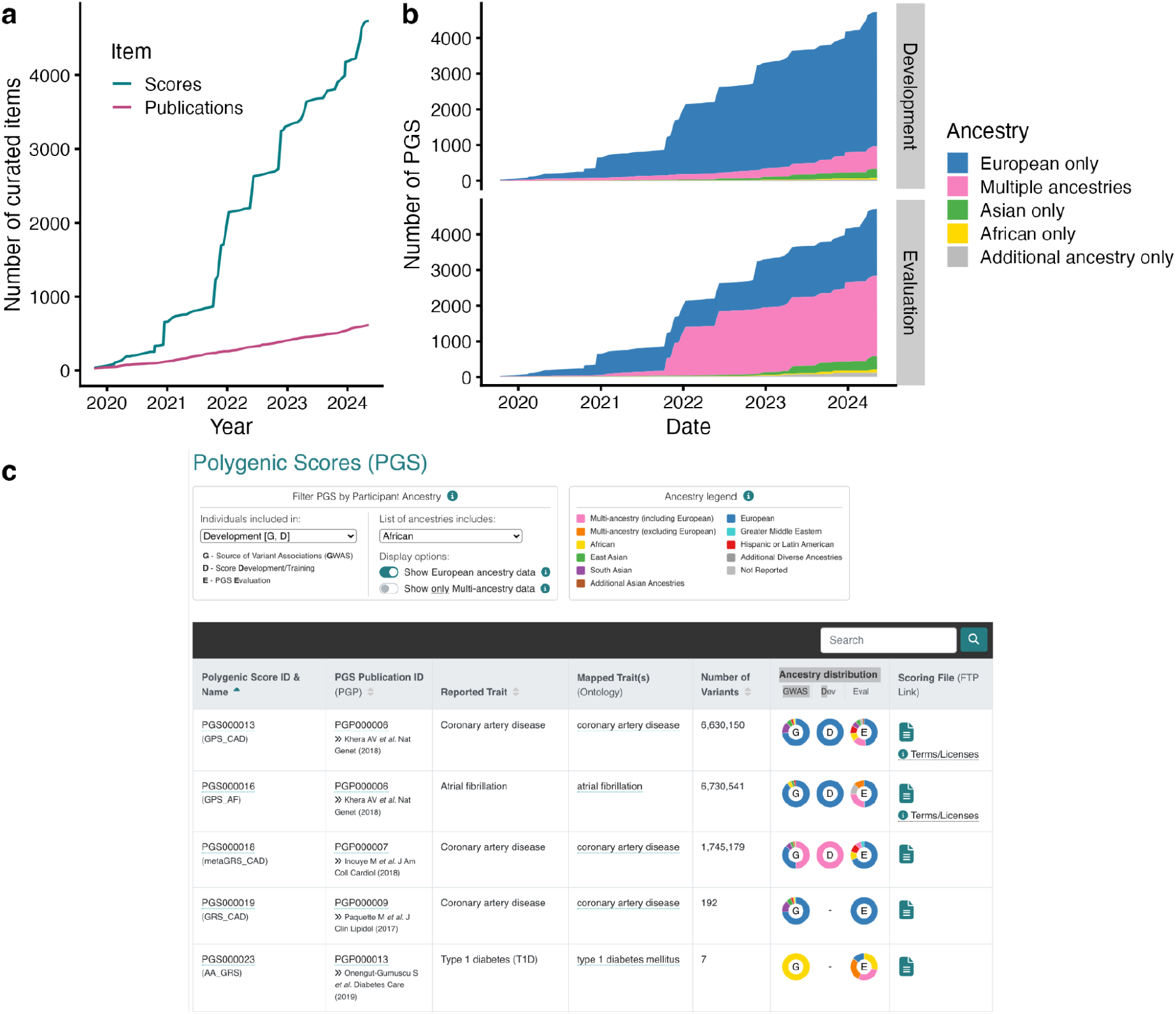
Data growth and improvement of the PGS Catalog. (**a**) Cumulative numbers of PGS (scores) and publications indexed in the PGS Catalog since inception (October 2019). **(b)** Cumulative numbers of PGS indexed in the Catalog since inception stratified by ancestry groups contributing to PGS development (i.e. GWAS and score development, top panel) or evaluation (bottom panel). Ancestry groups with lower numbers of PGS have been combined into larger groupings to visualize broader trends in single-ancestry and multi-ancestry data. **(c)** Example of ancestry filter present on every table with multiple PGS, the filter has been set to identify any PGS with African ancestry samples used for development (shown here are the first 5 of 348 PGS).

It is well documented that PGS developed using genetic data from participants of predominantly European ancestries have reduced predictive performance in populations of non-European ancestries^14^; however, some scores developed using multi-ancestry data or current methods have improved transferability.^7,15^ Thus, it is important to understand the ancestral makeup and diversity of the samples used to develop a PGS in order to assess whether it may be a useful genetic predictor for any new population. We developed a new interface for all PGS tables on the website that allows users to visualize and filter scores that include a specific ancestry group or multi-ancestry data at any stage of score development and evaluation (**Figure 1**). As an example, this can be used to identify scores with measured performance metrics in a specific ancestry group. The ancestral diversity of data in the PGS Catalog has also grown (**Figure 1**). The number of PGS in the Catalog being developed using multi-ancestry or non-European ancestry data is steadily increasing but remains low as a proportion of total data; moreover, the majority of multi-ancestry development samples are predominantly made up of European individuals. The majority of PGS in the Catalog are now being evaluated in multiple ancestry groups, illustrating the effectiveness of recent efforts to raise awareness amongst researchers as to ancestry biases and corresponding transferability issues for PGS.

### Tools for more accessible and reproducible PGS calculation

Even with consistently formatted data from the PGS Catalog, it remains challenging to calculate PGS (i.e. applying scoring files to new genomic data) as it requires interacting with multiple different software tools required for genotype data formatting, variant matching, and allelic scoring that can also differ across platforms. Few tools dedicated to calculation have been developed or implemented in a workflow manager (e.g. Nextflow or Snakemake) which limits their portability and scalability.^16^ Also critical to the use of PGS is the ability to report genetic distribution on an interpretable scale since many PGS have distributions that are confounded by ancestry.^17,18^ Currently, no standalone software exists to adjust polygenic scores across genetic ancestry groups using standard methods^19–21^. This is a key barrier to the equitable application of polygenic scores. A tool combining these features would increase reusability of PGS data and lower the barrier to entry of PGS analysis to users without specific training in bioinformatics, an important consideration to facilitate clinical studies. Additionally, given the sensitivity of human genetic data, solutions should easily allow users to bring “code to the data’’ without requiring data submission to web servers, which raises obvious governance issues.

To address these gaps, we developed the PGS Catalog Calculator (`pgsc_calc`), a reproducible workflow for PGS calculation, ancestry analysis, and adjustment of calculated scores. The workflow is optimized to enable the user to calculate PGS at biobank scale across diverse computing environments, including High Performance Compute clusters (HPCs), airlocked Trusted Research Environments (TREs), and cloud architectures. Users can specify any combination of scores from the PGS Catalog for calculation using score, publication, or trait identifiers, or supply their own custom PGS scoring files. PGS scoring results are aggregated and output into a results file along with a summary report.

The PGS Catalog Calculator is implemented in Nextflow, a popular workflow system used by many bioinformaticians.^22^ Nextflow orchestrates parallel computational analyses on multiple platforms and job schedulers (e.g. AWS, Google, SLURM) using the most common software containers for reproducibility (docker, singularity, conda), and has a community framework for open-source development (nf-core).^22,23^ The pipeline is developed and maintained by the PGS Catalog team under a permissive software license (Apache v2.0). The code is open-source and available on Github (https://github.com/PGScatalog/pgsc_calc), which also hosts the documentation (https://pgsc-calc.readthedocs.io/), an active issues tracker, and discussion forum to offer support and communicate with users. Contributions (pull requests) from other developers are welcomed.

An overview of the workflow is shown in **Figure 2** and the implementation is described in detail in the **Supplemental Note**. In detail, PGS Catalog Calculator takes an input of target genomes in most commonly-used genomics formats (variant call format [VCF], or PLINK 1 or 2 format^24^) and genome build information for PGS calculation. The input file(s) can either contain all variants, or be split to have one file per chromosome to enable parallelization and faster calculation. Scorefiles from the PGS Catalog are automatically downloaded and combined with any custom scoring files. Scoring file variants are matched to variants in the target genotypes, automatically handling typical but complex scenarios of multi-allelic variants, strand flips, and scores with only the effect allele with sensible default settings.

**Figure 2.**
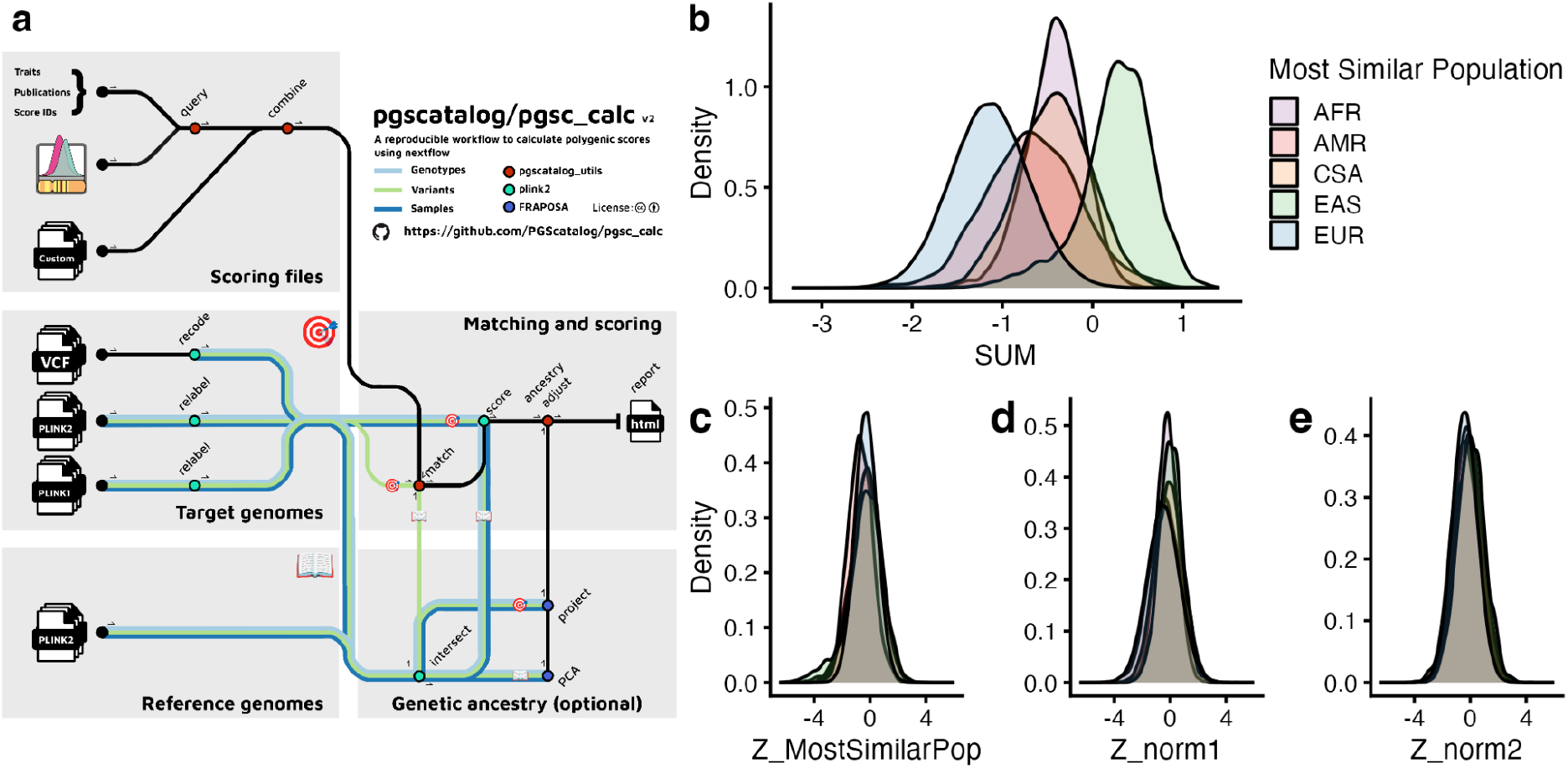
The PGS Catalog Calculator for reproducible PGS calculation with estimation and adjustment for genetic ancestry. (**a**) Visual summary of the PGS Catalog Calculator (pgsc_calc) pipeline, displaying the inputs, outputs, and data flows between modules. (b-e) To illustrate the utility of the pipeline we calculated PGS000018 (metaGRS_CAD_, 1,745,179 variants)^31^ applied in all ∼487,000 individuals with imputed genotypes in UK Biobank^32^ using pgsc_calc and ran the ancestry adjustments using HGDP+1kGP as a reference panel. (**b**) The PGS calculated as a weighted `SUM` of dosages has different distributions in different ancestry groups (grouped based on their Most Similar Population label from the reference panel: African [AFR], American [AMR], Central and South Asian [CSA], East Asian [EAS], and European [EUR] superpopulation ancestry). (**c-e**) Ancestry adjustment methods applied to UK Biobank data make PGS distributions more comparable across populations. The most similar ancestry group labels are used to normalize PGS using their most similar reference population distribution (`Z_MostSimilarPop`, **c**). Population labels are not used in continuous ancestry adjustment methods that adjust for mean (`Z_norm1`, **d**) and variance (`Z_norm2`, **e**) but are grouped in order to visualize the overlap after normalization.

Final scoring files are created, along with an auditable log summarizing the variants that have been excluded and why. PGS scoring is applied to genotypes in parallel, and the results are aggregated and output into a simple tabular results file along with a report that summarizes the scores calculated, their matching, and the PGS distributions (see **Supplemental Note** for details). The results file containing the calculated PGS can easily be combined with phenotype information for downstream PGS analyses.

A key challenge is that PGS are often on different scales that cannot be easily interpreted and have no natural cut-points. A common solution is to express genetic predisposition as a relative measure by standardizing the PGS calculation to the sample mean and standard deviation; however, this has the problem of being sensitive to the set of samples included in the specific run of PGS calculation. Importantly, PGS distributions can also be confounded by genetic ancestries (**Figure 2**), i.e. differences in PGS means and variances which arise from differential allele frequencies and linkage disequilibrium between populations^17,18^.

These differences do not necessarily correspond to differences in disease risk (e.g. changes in disease prevalence) or quantitative trait values between the populations; however, genetic ancestry is important for estimating relative risk.

The PGS Catalog Calculator implements common methods to normalize PGS in the context of genetic ancestry to report relative risk information using population reference panels, these include: comparison to a reference distribution of scores from a similar population^21^, and continuous PCA-based adjustments^19,20^ (see **Supplemental Note** and **Supplementary Figure 1** for overview). As default, the PGS Catalog Calculator uses the largest open dataset of globally-representative genotyped individuals from the Human Genome Diversity Panel and 1000 Genomes Project (HGDP+1kGP)^25^ as a reference panel, to visualize PGS distributions and adjust PGS using ancestry similarity and projections.

Methods for ancestry adjustment of PGS start by determining the overlap of variants and target genotypes, then creating a principal component analysis (PCA) of the reference panel and projecting individual(s) into the genetic ancestry space using the Online Augmentation, Decomposition and Procrustes (OADP) method to avoid issues of PCA shrinkage.^26^ The PCA projections are used to determine the population in the reference panel to which the individual is most similar. The population labels in the reference panel are not comprehensive or deterministic, thus samples from the target dataset are only described in terms of their genetic similarity to these populations. This avoids transfer of ancestry labels that may not reflect the new individuals.^27^ The relative PGS for each individual can then be calculated by comparing the calculated PGS to the distribution of PGS in the most similar population in the reference panel and reporting it as a percentile or Z-score (`Z_MostSimilarPop`, **Figure 2**).

When treating ancestry as a continuum (represented by loadings in PCA-space) or when an individual has an ancestry not in the reference panel, a relative PGS is also calculated without the use of population labels together with regression to remove the effects of genetic ancestry on PGS distributions. Using regression has the benefit of not assigning individuals to specific ancestry groups, which may be particularly problematic for empirical methods when an individual has an ancestry that is not represented within a reference panel or has recent admixed ancestry (custom reference panels can be used instead of the HGDP+1kGP default). The continuous methods work by correcting differences in mean PGS observed across ancestry groups (`Z_norm1`, **Figure 2**)^19^, and additionally correcting for the differences in the variance of the distributions (`Z_norm2`, **Figure 2**).^20^ These methods of adjusting PGS using continuous ancestry representations have been used to report genetic risk in clinical trials, with mean normalization (`Z_norm1`) being used in the GenoVA trial^11^, and variance normalization in eMERGE (`Z_norm2`).^28^ Despite the necessity of rescaling PGS to a more interpretable relative risk scale, these adjustment methods have thus far not been implemented in a reproducible package or workflow (see **Supplemental Note** for detailed comparison to other tools^24,29,30^). We will continue to optimize these adjustments within the PGS Catalog Calculator, and our open-source package and workflows allow for the addition of new methods as they are developed.

## Conclusions and future developments

As the research and clinical applications for polygenic scores continue to grow so too will the PGS Catalog. This has been particularly evident with the large growth in PGS and evaluations from diverse ancestry groups, allowing for the construction of new features that allow users to browse these data more easily. The PGS Catalog will continue to evaluate how to best represent ancestral diversity to increase genomic equity and in accordance with current best practices.^27,33^ In order to make PGS as accessible as possible we are committed to increasing the ease of data deposition into the PGS Catalog and are currently developing an automated deposition portal in collaboration with the GWAS Catalog^34^, our sibling resource, to make it easier for users to validate and upload data. Furthermore, we will continue to develop our documentation, training materials, and deliver workshops to support users and maximize the impact of open sharing of PGS.

To democratize secure PGS calculation and ancestry adjustment, we developed the PGS Catalog Calculator. This software tool provides advantages to the user, notably the implementation of methods to ensure PGS distributions are consistent among genetic ancestry groups, which represents a barrier to the equitable application of PGS. The calculated scores are all forms of relative risk depending on the method used and provided in a simple format to allow users to integrate these results into other steps of genetic prediction studies (e.g. measuring PGS-trait associations, absolute risk predictions) if they choose. The software has been successfully deployed to various computing environments including university and cloud HPCs, airlocked TREs, and integrated into online web server/platforms and has an active user community on our Github. Future versions will further improve ancestry estimation and normalization, and implement newer methods for PGS calculation as they are developed. The current PGS Catalog Calculator is optimized to calculate PGS on imputed genotypes derived from genotyping array data; however, a common user request is to support whole-genome sequencing data (unimputed VCFs). In future, tools to support WGS data will be developed, thus ensuring that PGS can be equitably and reproducibly applied to all genomic data. Taken together, the PGS Catalog now provides an integrated suite of tools to support the use of PGS in clinical studies and biomedical research more broadly.

## Supporting information

Supplemental Note (includes Supplementary Figures 1-6, and Supplementary Tables 1-2)

## Data & code availability statement

Data in the PGS Catalog is distributed according to EBI’s terms of use (https://www.ebi.ac.uk/about/terms-of-use) or specifically marked with open-access licenses supplied by authors. All code for the PGS Catalog project and calculator are hosted on GitHub under the PGS Catalog organization (https://github.com/PGScatalog/) and released under an Apache v2 license. The processed reference panels 1kGP and 1kGP+HGDP are available from our FTP server (https://ftp.ebi.ac.uk/pub/databases/spot/pgs/resources/). UK Biobank data can be accessed via application, the data for this publication was accessed as part of projects 49978 and 78537.

## Acknowledgements

We wish to thank all the authors of publications in the PGS Catalog for making their data available and indexable in our database, and all those who responded to our inquiries and requests for data. We would also like to thank users of the workflow, especially those that have contributed to bug reports, feature requests, or discussions.

This work was supported by core funding from the British Heart Foundation [RG/18/13/33946, RG/F/23/110103], National Human Genome Research Institute of the National Institutes of Health grant [1U24HG012542-01 to H.P. M.I.], NIHR Cambridge Biomedical Research Centre [BRC-1215–20014, NIHR203312], Cambridge BHF Centre of Research Excellence [RE/18/1/34212, RE/18/1/34212], BHF Chair Award [CH/12/2/29428], European Union’s Horizon 2020 research and innovation programme [under grant agreement No 101016775 INTERVENE to H.P. M.I.and B.W.], as well as by Health Data Research UK, which is funded by the UK Medical Research Council, Engineering and Physical Sciences Research Council, Economic and Social Research Council, Department of Health and Social Care (England), Chief Scientist Office of the Scottish Government Health and Social Care Directorates, Health and Social Care Research and Development Division (Welsh Government), Public Health Agency (Northern Ireland), British Heart Foundation and Wellcome.The views expressed are those of the authors and not necessarily those of the NHS, the NIHR or the Department of Health and Social Care. This work was performed using computing resources provided by the Cambridge Service for Data Driven Discovery (CSD3) operated by the University of Cambridge Research Computing Service (www.csd3.cam.ac.uk), provided by Dell EMC and Intel using Tier-2 funding from the Engineering and Physical Sciences Research Council (capital grant EP/T022159/1), and DiRAC funding from the Science and Technology Facilities Council (www.dirac.ac.uk). The authors wish to acknowledge CSC—IT Center for Science, Finland, for providing computing resources.

M.I. is supported by the Munz Chair of Cardiovascular Prediction and Prevention and the UK Economic and Social Research 878 Council (ES/T013192/1). H.P. is supported by European Molecular Biology Laboratory Core Funds.

