## Supplemental Note (includes Supplementary Figures 1-6, and Supplementary Tables 1-2) for "The Polygenic Score Catalog: new functionality and tools to enable FAIR research"

#### Creating harmonized scoring files to facilitate easier PGS calculation

PGS Scoring Files in the Catalog are currently provided in a consistent format with standardized column names and data types, along with information about the genome build given by authors (see https://www.pgscatalog.org/downloads/#dl_scoring_files for schema). The variant-level information in these PGS is often heterogeneously described and may lack chromosome/position information, contain a mix of positions and/or rsIDs, or be mapped to a genome build different from target genotypes. To make make variant matching and PGS calculation easier we developed software to harmonize scoring files into a new scoring file for each commonly used genome build (currently, GRCh37/hg19 and GRCh38/hg38) by ensuring variant information (chromosome name and base pair position) and variant identifiers (updated rsID) are present.

The generation of these harmonized files is done by using the `pgs-harmonizer` tool (https://github.com/PGScatalog/pgs-harmonizer) for each new or updated scoring file as part of the PGS Catalog data release process. It is based on the Open Targets and GWAS Catalog Summary Statistics harmonizer pipelines (https://github.com/EBISPOT/gwas-sumstats-harmoniser). To harmonize the variant positions the `pgs-harmonizer` performs the following tasks:

- Mapping rsIDs to chromosomal positions using Ensembl (VCF files and REST APIs) on GRCh37 and GRCh38. Currently, the PGS Catalog maps data using Ensembl version 105.
- Liftover - if only chromosomal positions are provided we map chromosomal positions across builds using the UCSC liftover tools via the Python library `pyliftover` (https://github.com/konstantint/pyliftover). This only occurs when generating a Scoring file on a different genome build (the author-supplied positions are otherwise reported).

The resultant files create new columns (see https://www.pgscatalog.org/downloads/#hm_pos_columns for schema), indicating the source of the variant annotation (hm_source), as well as consistently annotated chromosome (hm_chr) / position (hm_pos), and rsID (hm_rsID) which can be used to match scoring file variants in target genomes in combination with the alleles (effect_allele, and other_allele).

#### Development and implementation of the PGS Catalog Calculator (`pgsc_calc`) within Nextflow and Python

Each process within the workflow is contained in a module that ideally contains a single software tool, which are provided as Docker and Singularity containers via the GitHub container registry. To support environments without containers, Anaconda environment files are provided. The use of containers ensures reproducibility, and portability of the software - this allows the pipeline to run at the location of the data, which is essential as human genomes are sensitive data and many users will be unable to move their datasets to other computing environments. Multiple modules are organized into a subworkflow that performs a specific task, and the current `pgsc_calc` pipeline includes subworkflows for input checks, data harmonization, variant matching/scorefile creation, PGS calculation, and optional analyses involving reference panels and genetic ancestry.

The pipeline uses synthetic data for automated, continuous integration testing (in Docker and Singularity environments), which is deployed using GitHub Actions. The datasets include:

- a small subset variants from the CINECA synthetic European cohort for unit testing (EGA accession: EGAD00001006673)
- the small HAPNEST[1] dataset for end to end tests of ancestry estimation (BioStudies accession: S-BSST936)

Our Nextflow implementation supports automatic parallelization of many steps of the workflow, handling the most common scenario where the genotyping data is split by chromosome. During development we made efforts to ensure each stage of the workflow can scale to biobank sized data. For example, we regularly run the workflow using hundreds of scores on the UK Biobank, which contains ~500,000 samples. These analyses typically run on a HPC and require up to 64GB of RAM and 4 CPU. The runtime of the pipeline is primarily determined by the number of unique variants being scored and the number of samples - therefore it is most efficient to run the workflow once with multiple scores instead of running the workflow many times with a single score.

The `pgsc_calc` pipeline uses plink2 internally for genotype data harmonization and PGS calculation.[2] plink2 is a standard genomics toolkit, and we reuse these features to avoid duplicating existing work.[2] However, we have developed independent software packages to automate and improve many aspects of scoring genotypes with plink2 (see **Supplementary Table 1**). `pgscatalog_utils` (https://github.com/PGScatalog/pygscatalog, https://pypi.org/project/pgscatalog-utils/) is a python package that offers a set of tools for working with the PGS Catalog API, polygenic scoring models, and calculated scores. `fraposa_pgsc` (https://github.com/PGScatalog/fraposa_pgsc) is an improved fork of FRAPOSA[3], a python package which implements a more accurate method for projecting new samples using existing genetic principle components that removes the effects of shrinkage. We improved the original FRAPOSA software by adding extra checks to ensure the genotype and allele orientation is correct between the reference and target samples. All the component software packages within the workflow are open source and publicly available, and can also be used as standalone tools in other settings (**Supplementary Table 1**).

**Supplementary Table 1. Workflow component dependencies.**

| Workflow component | plink2 | pgscatalog_utils | fraposa_pgsc |
|---|---|---|---|
| PGS Catalog API integration to download polygenic score models | | ✅ | |

| | | | |
|---|---|---|---|
| Genotype data harmonization | ✅ | | |
| Merging PGS scoring files for matching | | ✅ | |
| Variant matching between scoring file and target genotypes | | ✅ | |
| PGS scoring | ✅ | | |
| Calculating PGS by aggregating data split across chromosomes | | ✅ | |
| Derivation of genetic PCA space & projection of new samples | | | ✅ |
| Ancestry similarly analysis and PGS adjustment | | ✅ | |

#### Detailed description of `pgsc_calc` pipeline

Here we describe step-by-step the methods used to calculate PGS within the PGS Catalog Calculator (`pgsc_calc v2.0.0`, see **Figure 2** for a visual overview):

##### Genotype data inputs & harmonization

PGS Catalog Calculator takes an input of target genomes and build information for PGS calculation in most commonly-used genomics formats (variant call format [VCF], or PLINK 1 or 2 format[2]) and assumes the data has been imputed using an reference panel to increase the likelihood that PGS variants are matched. The workflow begins by recoding genotypes into PLINK2 compressed binary format and relabelling variant identifiers in a consistent way to make variant matching easier. VCFs must be recoded into a triplet of files: a variant information file, a genotype data file, and a sample information file. PLINK1 files are backwards compatible with PLINK2, so only new variant information files are created when working with this type of data to reduce I/O operations. Sample information is not currently used by the workflow, but may be in the future (e.g. to enable sex checking).

##### Scoring files: PGS Catalog integration & custom inputs

We integrate with the PGS Catalog API (https://www.pgscatalog.org/rest/) to download polygenic scoring files using search terms like Polygenic Score ID (`--pgs_id`), Publication ID (`--pgp_id`), or Trait ontology ID (`--trait_efo`). Harmonized scoring files that contain genomic coordinates which match input genome build (GRCh37 or GRCh38) and complete allele information are fetched according to workflow parameters.

Custom user defined scoring files are also supported with the `-scorefile` parameter. Custom files should be formatted in the PGS Catalog v2 specification (https://www.pgscatalog.org/downloads/#scoring_header) and minimally contain a header with the build information, and columns with a chromosomal position, effect allele, and effect weight - a non-effect allele is also recommended to improve matching accuracy. Scoring files with builds that do not match the target genotypes will automatically have positions lifted to the new build using the `pyliftover` (https://github.com/konstantint/pyliftover) tool.

All scoring files are merged into a common representation for input into the variant matching steps.

#### Variant matching

Variant matching is critical to faithfully reproduce scores deposited in the PGS Catalog. It's difficult to unambiguously match a variant described in a scoring file against genotypes: variant identifiers (e.g. dbSNP rsID) are not stable or unique identifiers, genomic coordinates change across different genome builds, and matching effect alleles can be technically challenging because of allele flips and strand mismatches. The PGS Catalog scoring file standard and genome build-harmonized data uploads simplify this process by making structured data available that includes important information like effect allele and other allele. The variant matching procedure is as follows:

1. Multiallelic variants in the target genomes are split so that all REF/ALT combinations can be matched (optional and configurable by user)
2. Identify all potential matches where chromosome and position must match and:
    a. [effect_allele, other_allele] in scoring file == [REF, ALT] in target genomes
    b. [effect_allele, other_allele] in scoring file == [ALT, REF] in target genomes
    c. Complemented [effect_allele, other_allele] in scoring file == [REF, ALT] in target genomes (optional, configurable by user parameter)
    d. Complemented [effect_allele, other_allele] in scoring file == [ALT, REF] in target genomes (optional, configurable by user parameter)
    e. If other allele information is missing, drop this data (and its corresponding field in the target genome) from comparison
3. Label match candidates with flags to describe variants that are:
    a. Strand **ambiguous**,e.g. A/T, C/G SNPs
    b. **Multiallelic** variants
    c. **Duplicated**, same scoring file variant matches multiple variants
    d. **Flipped** strand, the complement of the variant is present in target genomes
    e. **Present in an external list of variants eligible for matching** (optional, used during ancestry adjustment to calculate scores on a consistent set of variants)
4. For each variant in the original scoring file, select the best possible match after:
    a. Ranking match candidates in order of matching strategies 2.a-d (*a is preferred*)
    b. Excluding match candidates according to user parameters, defaults:
        i. No multiallelic variant matches (can be altered using `--keep_multiallelic`)
        ii. No ambiguous variant matches (can be altered using `--keep_ambiguous`)
5. Comparing the best match candidates to the original scoring file, calculate the percentage of variants that are matched and remain after exclusions.
    a. If the overlap for a score is below 75% (user configurable with `--min_overlap`), drop the score from step 7 but continue
    b. If all unique scores are below 75%, stop matching and raise an error
6. From the union of match candidates and missing matches, create an auditable log with one variant per row that includes:
    a. Scoring file variant information

b. Target genome variant information
c. Match metadata including labels
7. Pivot best match candidates to wide format, so that variant IDs with multiple matched effect weights are transformed to have multiple columns for parallel score calculation
   a. Missing effect weights generated by pivoting are filled with 0
   b. Write this data to a compressed text file in plink2 scoring file format

A default minimum threshold is set - at least 75% of variants in a scoring file must be successfully matched to genotyped sites in the target file before calculation can proceed - to reduce the chance of polygenic score misapplication (`--min_overlap` parameter can be adjusted). After matching, new intermediate scoring files are written to enable efficient polygenic score calculation in parallel, and an auditable log is written for the union of variants across all scoring files.

#### PGS calculation

PGS are calculated as linear sums of the effects of *m* variants, based on the the effect weight in the scoring file ($\beta$):

$$PGS_i = \sum_{j}^{m} x_{ij}\beta_j$$

where $x_{ij}$ is the genotype for the $i^{th}$ individual and $j^{th}$ SNP (usually encoded as 0, 1 or 2 for the effect allele dosage). Scoring files produced by the variant matching process are applied to genotypes using `plink2 --score`. Duplicate variants with different effect alleles are automatically split into separate files to overcome plink limitations. Scoring is automatically parallelized across chromosomes if the input genomes are split. If the reference panel is used, then allelic frequencies are automatically calculated from the reference panel and loaded to impute missing genotypes. This is particularly useful when applying PGS to low sample sizes (fewer than 50) where in-sample imputation is more likely to be unreliable (see plink documentation). Computing multiple scores in parallel adds very little computational resources (the pipeline computation time scales with the number of variants in the scoring files, not the number of scores). It's far more efficient to run the workflow once with multiple scores than to run the workflow once for each score (see below). Calculated scores are automatically aggregated once scoring has finished to simplify downstream applications of PGS.

#### Ancestry adjustment (optional)

##### Derivation of population reference panels for use in PGS Catalog Calculator

To facilitate the analysis of PGS in the context of ancestry, the `pgsc_calc` pipeline distributes and can create reference panels of individuals with population descriptors (e.g. genetic ancestry groups). The pipeline by default uses a merged variant callset of the 1kGP and Human Genome Diversity project (HGDP) samples for build GRCh38 release in gnomAD (v3.1.2), adding additional diversity to the 1kGP data, specifically Middle Eastern and Oceanian ancestry individuals.[4] To prepare the HGDP+1kGP data for use in the `pgsc_calc` pipeline we filtered the data to all high-quality individuals and variants passing gnomAD QC filters, additionally filtering to variants with a minor allele count greater than 10, and converting plink2 pgen format. To provide a version of the new panel in GRCh37 we applied the same filters and used the GATK[5] liftoverVCF tool, again converting the data to plink2 pgen format. The `pgsc_calc` pipeline can then be run in combination with these reference panels to calculate PGS as a relative risk measure similar to individuals of a similar genetic ancestry.

##### Description of ancestry workflow

Variants in the reference panel are first intersected with the target genomes. This intersection is used to match against scoring files so that both the target genomes and reference panels use the same set of variants to calculate PGS for consistency. Allelic frequencies are calculated in the reference panel and loaded while scoring target genomes to impute missing genotypes.

A PCA is derived using fraposa-pgsc on the reference panel, filtered to unrelated samples with low genotype missingness (<10%) with standard filters for variant-level QC (SNPs in Hardy–Weinberg equilibrium [p > 1e-04] that are bi-allelic and non-ambiguous, with low missingness [<10%], and minor allele frequency [MAF > 5%]). LD-pruning is then applied to the variants and sample passing these checks (r2 threshold = 0.05), excluding complex regions with high LD (e.g. MHC).[6] The LD-pruned variants of the unrelated samples passing QC are then used to define the PCA space of the reference panel (default: 10 PCs) using FRAPOSA.[3] The PCA of the reference panel (variant-PC loadings, and reference sample projections) are then used to determine the placement of the target samples in the PCA space using projection. Naive projection (using loadings) is prone to shrinkage which biases the projection of individuals towards the null of an existing space, which would introduce errors into PCA-loading based adjustments of PGS. For a less biased projection of individuals into the reference panel PCA space we use the online augmentation, decomposition and Procrustes (OADP) method of the FRAPOSA package.[3] We chose to implement PCA-based projection over derivation of the PCA space on a merged target and reference dataset to ensure that the composition of the target doesn't impact the structure of the PCA. This is implemented in the FRAPOSA_OADP module. We note that the quality of the PCA and projection should be visually assessed in the report - further optimizations and QC steps will be added to avoid manual inspection in the future.

The calculated PGS (SUM), reference panel PCA, and target sample projection into the PCA space are supplied to a script that performs the analyses needed to adjust the PGS for genetic ancestry. This functionality is implemented within our pgscatalog_utils package, and is comprised of two steps: genetic similarity analysis and PGS adjustment (see **Supplementary Figure 1** for overview of methods).

Performing empirical PGS adjustments requires comparing an individual's PGS to a distribution of scores in a reference population. To determine a suitable reference population we perform a genetic similarity analysis using the PCA projections to determine the most similar population for each sample. By default this is done by fitting a RandomForest classifier to predict reference panel population assignments using the PCA-loadings (default: 10 PCs) and then applying the classifier to the target samples to identify the most genetically similar population in the reference panel (e.g. highest-probability).[4,7] Alternatively, the Mahalanobis distance between each individual and each reference population can be calculated and used to identify the most similar reference population (minimum distance).[8]

The probability of membership for each reference population and most similar population assignments are recorded and output for all methods, heuristic p-value thresholds are used to determine low-confidence similarity labels (0.5 for RandomForest, and 1e-10 to determine outliers via a Mahalanobis converted into a p-value using the Chi-squared distribution).

The empirical PGS adjustments (`percentile_MostSimilarPop`, `Z_MostSimilarPop`) are then performed by combining the results of the genetic similarity analysis. The relative PGS for each individual is calculated by comparing the calculated PGS to the distribution of PGS in the most similar population in the reference panel and reporting it as a percentile (output column: `percentile_MostSimilarPop`) or as a Z-score (output column: `Z_MostSimilarPop`).

To perform the PCA-based adjustments (`Z_norm1`, `Z_norm2`) only the PGS and PCA-loadings are used using regression to correct for mean, or mean and variance shifts that are correlated with genetic ancestry. By default these methods use the first 4 components of genetic ancestry, but this setting can be changed in the pipeline interface. The first method (originally proposed by Khera et al.[9]) to adjust for differences in the means of PGS distributions across ancestries by fitting a linear regression of PGS values based on PCA-loadings using unrelated individuals of the reference panel:

$$PGS \sim PC_{1-4} \text{ (Eq. 1)}$$

and the observed standard deviation of the PGS in the reference panel ($\sigma_{ReferencePanel}$). The regression is then used to calculate the ancestry-predicted PGS ($PGS_{Pred}$) based on the target sample's PCA projections and the normalized PGS is calculated as:

$$PGS_{Znorm1} = (PGS_{SUM} - PGS_{Pred})/\sigma_{ReferencePanel}$$

This achieves PGS distributions that are approximately centered at 0 for each genetic ancestry group (output column: `Z_norm1`), while not relying on any population labels during model fitting. The first method (`Z_norm1`) has the result of normalizing the first moment of the PGS distribution (mean); however, the second moment of the PGS distribution (variance) can also differ between ancestry groups.[10] A second regression of the PCA-loadings on the squared residuals (difference of the PGS and the predicted PGS)

$$(PGS_{SUM} - PGS_{Pred})^2 \sim PC_{1-4} \text{ (Eq. 2)}$$

can be fit using a Gamma regression in the same unrelated individuals of the reference panel to estimate a predicted standard deviation ($\sigma_{Pred}$) based on genetic ancestry, as was proposed by Khan *et al.* and implemented within the eMERGE GIRA.[10,11] The intercepts and coefficients from equations 1 and 2 are used and initializations to full likelihood model (originally implemented by Linder *et al.* in R https://github.com/broadinstitute/palantir-workflows/blob/v0.10/ImputationPipeline/ScoringTasks.wdl and re-implemented in Python within pgscatalog_utils)[11]:

$$PGS_{Znorm2} = (PGS_{SUM} - PGS_{Pred})/\sigma_{Pred}$$

This yields a new estimate of relative risk (output column: `Z_norm2`) where the variance of the PGS distribution is more equal across ancestry groups and approximately 1.

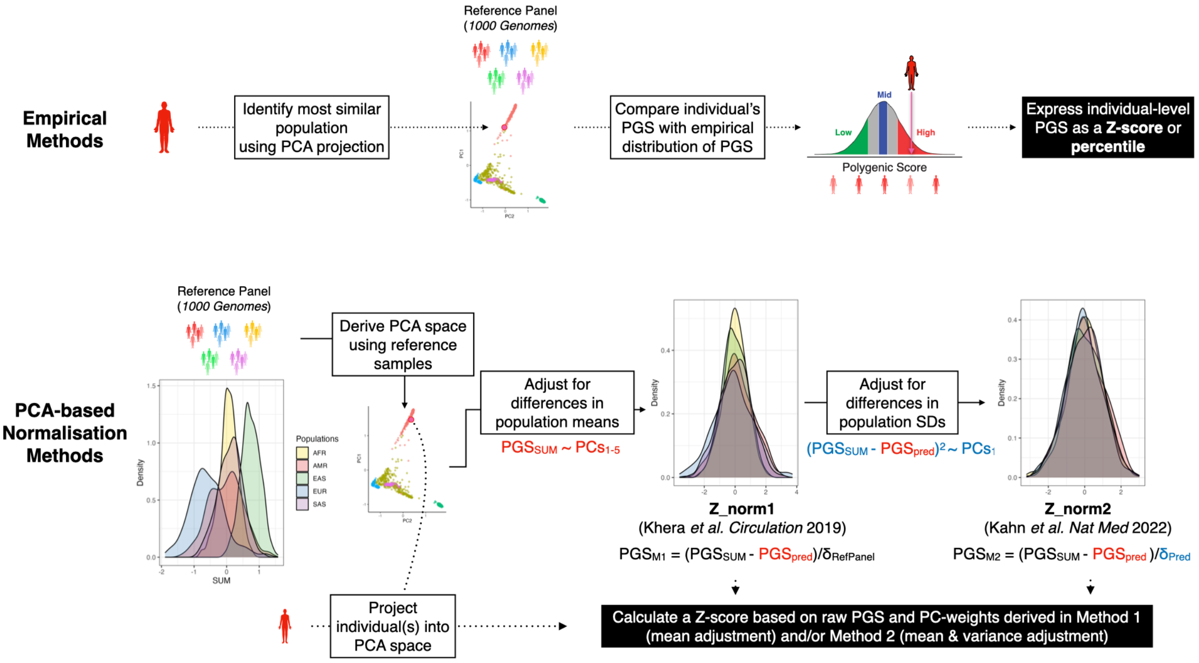


**Supplementary Figure 1. Schematic figure detailing empirical and PCA-based methods for contextualizing or adjusting PGS with genetic ancestry.** Data is for the normalization of PGS000018 (metaGRS$_{CAD}$) when applying `pgsc_calc --run_ancestry` to HGDP genotypes using 1kGP as a reference panel.

#### Description of `pgsc_calc` outputs

The outputs of `pgsc_calc` are described in our online documentation (https://pgsc-calc.readthedocs.io/en/latest/explanation/output.html), and we provide a brief overview here.

##### Aggregated PGS file

Calculated scores are stored in a gzipped-text space-delimited text file called `[sampleset]_pgs.txt.gz`. The data is presented in long form where each PGS for an individual is presented on a separate row (length = n_samples*n_pgs), and there will be at least four columns with the following headers (`sampleset`, `IID`, `PGS`, `SUM`). If the pipeline was run using ancestry information (`--run_ancesty`) the columns describing the ancestry adjustments that were calculated will also be present (`percentile_MostSimilarPop`, `Z_MostSimilarPop`, `Z_norm1`, `Z_norm2`). The aggregated file can be easily read into any analysis software (e.g. R/python) and linked to other datasets based on the identifiers in the genotyping data for downstream analysis.

##### Report

A summary report is provided for each successful run of the pipeline (`report.html`). The report can be opened in any web browser and contains useful information about the PGS that were applied, how well the variants in your target dataset match with the reference panel and scoring files, a summary of the computed genetic ancestry data, and some simple graphs displaying the distribution of scores in your dataset(s) as a density plot (**Supplementary Figures 2-3 and 5**). Some of the sections are only displayed when the ancestry analyses have been performed (**Supplementary Figure 4**).

### PGS Catalog Calculator (`pgsc_calc`) report

AUTHOR
PGS Catalog Calculator (`pgsc_calc`)

PUBLISHED
May 27, 2024

Table of contents


- Workflow metadata
  - Command
  - Version
- Scoring file metadata
- Variant matching
- Genetic Ancestry
- Population similarity summary
- Scores
- Citations


ⓘ **Note**

See the online documentation for additional explanation of the terms and data presented in this report.

#### Workflow metadata

##### Command

▶ Code

```
$ nextflow run pgscatalog/pgsc_calc -r v2.0.0-alpha.5 -latest -profile docker,arm \
    --max_cpus 4 --max_memory 32GB --input samples_splitHGDP.csv --target_build \
    GRCh38 --run_ancestry ./reference/pgsc_1000G_v1.tar.zst --genotypes_cache \
    cache_genotypes --pgs_id PGS000018 --outdir results18_v5 -w work18 -resume
```

##### Version

```
2.0.0-alpha.5
```

#### Scoring file metadata

##### Scoring file summary

▶ Code

CSV

Search:

| Scoring file | Polygenic Score ID | Publication | Traits | Number of variants | Genome build |
|---|---|---|---|---|---|
| PGS000018 hmPOS GRCh38 | PGS000018 metaGRS_CAD | PGP000007 Inouye M et al. J Am Coll Cardiol (2018). doi:10.1016/j.jacc.2018.07.079 | Reported trait: Coronary artery disease Mapped trait(s): coronary artery disease | 1745180 | Original build: GRCh37 Harmonised build: GRCh38 |

Showing 1 to 1 of 1 entries

Previous 1 Next

**Supplemental Figure 2. Example of pgsc_calc header.** First section of the report reproduces the nextflow command, and scoring file metadata (imported from the PGS Catalog for each PGS ID) describing the scoring files that were applied to the sampleset(s)

### Variant matching

#### Parameters

▶ Code

```
keep_multiallelic: false
keep_ambiguous   : false
min_overlap      : 0.75
```

#### Reference matching summary

Show 10 entries Search:

| | reference | n target | N variants in panel | n (matched) | % matched |
|---|---|---|---|---|---|
| 1 | 1000G | 83722464 | 73649067 | 40575977 | 55.09 |

Showing 1 to 1 of 1 entries Previous 1 Next

#### Summary

▶ Code

▶ Code

CSV Search:

| Sampleset | Scoring file | Number of variants | Passed matching | Match % | Tot matche |
|---|---|---|---|---|---|
| HGDP | PGS000018_hmPOS_GRCh38 | 1745179 | true | 97.1 | 1695 |

Showing 1 to 1 of 1 entries Previous 1 Next

#### Detailed log

CSV Search:

| Sampleset | Scoring file | Match type | Variant in reference panel | Ambiguous | Multiallelic | Matches strand flip | Multipl potenti matche |
|---|---|---|---|---|---|---|---|
| HGDP | PGS000018 hmPOS GRCh38 | matched | true | false | false | false | false |
| HGDP | PGS000018 hmPOS GRCh38 | matched | true | false | false | true | false |
| HGDP | PGS000018 hmPOS GRCh38 | excluded | false | false | false | false | false |
| HGDP | PGS000018 hmPOS GRCh38 | excluded | false | false | false | true | false |

**Supplemental Figure 3. Example of variant matching summaries in the pgsc_calc report.** The first table describes the number of variants in the target dataset that overlap with the reference panel (only present with `--run_ancestry`). The second table provides a summary of the number and percentage of variants within each score that have been matched, and whether that score passed the `--min_overlap` threshold (Passed Matching column) for calculation. The third table provides a more detailed summary of variant matches broken down by types of variants (e.g., strand ambiguous, multiallelic, duplicates) for matched, excluded, and unmatched variants.

#### Genetic Ancestry

```
# A tibble: 6 × 22
  sampleset IID          PC1   PC2   PC3    PC4    PC5   PC6    PC7    PC8    PC9
  <chr>     <chr>      <dbl> <dbl> <dbl>  <dbl>  <dbl> <dbl>  <dbl>  <dbl>  <dbl>
1 HGDP      HGDP00001 -18.5 -31.2 -19.7 -2.40   -0.761  5.61  0.973  1.34   2.56
2 HGDP      HGDP00003 -18.8 -29.9 -20.7 -0.866 -0.304  7.31 -0.186  0.350  5.56
3 HGDP      HGDP00005 -18.4 -29.7 -19.1 -4.00   -2.59   5.66  2.67   1.62  -0.907
4 HGDP      HGDP00007 -19.1 -32.1 -23.8 -0.913 -1.43   7.88 -0.120 -1.00   2.32
5 HGDP      HGDP00009 -18.2 -30.8 -21.5 -1.09   -0.721  6.75 -0.241 -0.931  1.09
6 HGDP      HGDP00011 -20.1 -32.1 -21.9 -0.693  0.879  6.62 -0.310  1.42   2.92
# ℹ 11 more variables: PC10 <dbl>, Unrelated <lgl>, RF_P_AFR <dbl>,
#   RF_P_AMR <dbl>, RF_P_EAS <dbl>, RF_P_EUR <dbl>, RF_P_SAS <dbl>,
#   MostSimilarPop <chr>, MostSimilarPop_LowConfidence <lgl>, REFERENCE <lgl>,
#   SuperPop <chr>
```

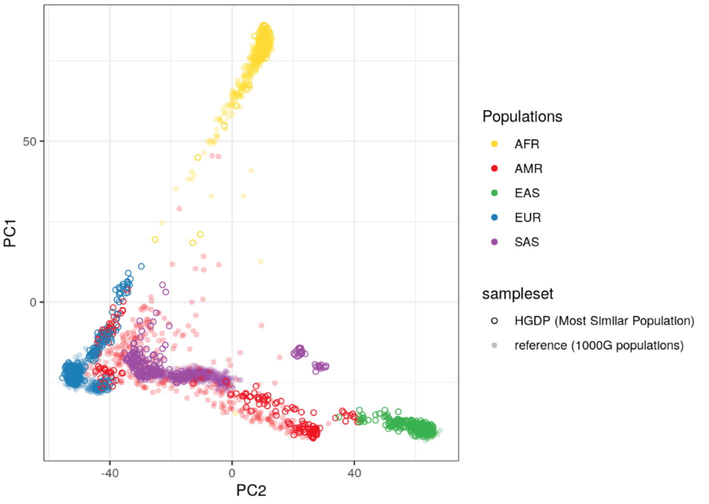


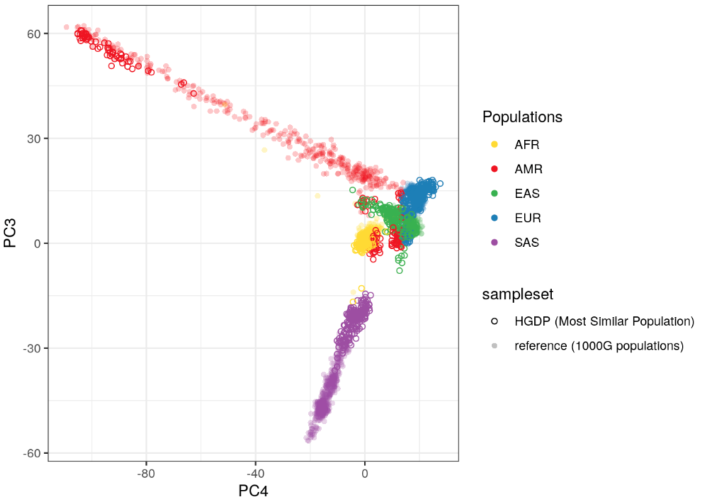


**Supplemental Figure 4. Visualization of genetic ancestry analysis within the report.** A a snippet of the `[sampleset]_popsimilarity.txt.gz` is followed by PCA plots for the first 6 PCs, where the target samples are colored according to the population that they are most similar to in the reference panel.

### Scores

```
[1] 3504
```

**Success**

- All requested scores were calculated successfully

1 scores for 3504 samples processed.

#### Score data

##### Score extract

**Note**

Below is a summary of the aggregated scores, which might be useful for debugging. See here for an explanation of plink2 column names

```
# A tibble: 3,504 × 8
   sampleset IID        PGS                      SUM Z_MostSimilarPop Z_norm1 Z_norm2
   <chr>     <chr>      <chr>                  <dbl>            <dbl>   <dbl>   <dbl>
 1 HGDP      HGDP00001 PGS000018_hmPOS_…  0.166            0.158   1.05    0.916
 2 HGDP      HGDP00003 PGS000018_hmPOS_… -0.658           -2.03   -1.18   -1.05
 3 HGDP      HGDP00005 PGS000018_hmPOS_… -0.279           -1.02   -0.196  -0.183
 4 HGDP      HGDP00007 PGS000018_hmPOS_… -0.645           -1.99   -1.09   -0.963
 5 HGDP      HGDP00009 PGS000018_hmPOS_…  0.183            0.202   1.10    0.955
 6 HGDP      HGDP00011 PGS000018_hmPOS_…  0.158            0.138   1.07    0.917
 7 HGDP      HGDP00013 PGS000018_hmPOS_… -0.122           -0.606   0.125   0.114
 8 HGDP      HGDP00015 PGS000018_hmPOS_… -0.424           -1.41   -0.490  -0.437
 9 HGDP      HGDP00017 PGS000018_hmPOS_…  0.164            0.153   1.09    0.941
10 HGDP      HGDP00019 PGS000018_hmPOS_… -1.41            -4.02   -3.21   -2.84
# ℹ 3,494 more rows
# ℹ 1 more variable: percentile_MostSimilarPop <dbl>
```

##### Density plot(s)

**Note**

The summary density plots show up to six scoring files

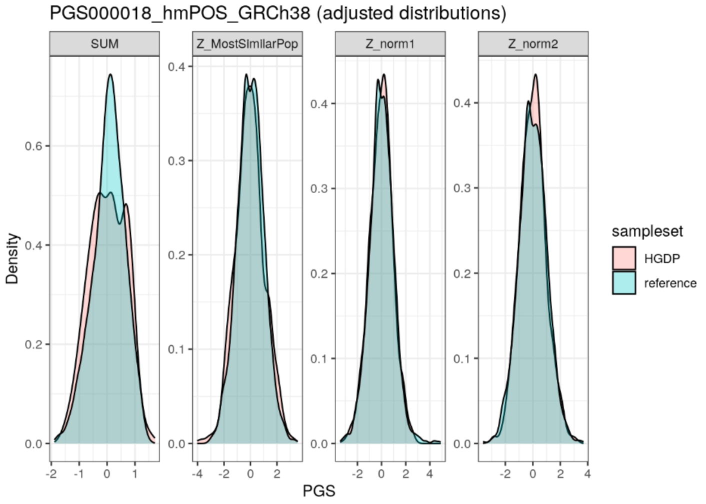


**Supplemental Figure 5. Example of the `[sampleset]_pgs.txt.gz` table and plots of PGS distributions.** A snippet of the PGS results file is displayed along with a visual display of the PGS distribution for a set of example score(s) for each method of PGS adjustment.

##### Population similarity summary

A second gzipped-text space-delimited text file called `[sampleset]_popsimilarity.txt.gz` will also be output, describing the analysis of the target samples in relation to the reference panel and ancestry labels. The file contains information about the PCA projection, genetic similarity analysis (probabilities of being from each reference panel population), and information about the reference panel samples (relatedness, population label). These data can be useful for stratifying analyses by most similar ancestry group, assessing PGS adjustments, or as a source of PCs for use in phenotype-PGS modeling.

#### Application of `pgsc_calc` to HGDP and UK Biobank data: scalability and performance

The PGS Catalog Calculator has supported biobank scale target genomes since v1.3.0 (https://doi.org/10.5281/zenodo.7342886). Each process in the workflow was profiled with a range of input data to gather statistics about CPU and memory usage. From these statistics we determined the main factor that influences scaling is the number of variants in the union of all scoring files and the variant density of the target genomes (see **Supplementary Figure 6**). Native support for parallelized score calculation means that running the workflow iteratively for each score is very inefficient compared to a single multi-score calculation. The number of distinct PGS being calculated has minimal impact on workflow runtime and resource usage compared with the total number of variants in the scoring file union.

The Nextflow dataflow model supports implicit parallelization. Tasks are automatically distributed to workers for each chromosome during the calculation process. Scoring jobs for each chromosome are also parallelised when required, such as when processing variants with recessive or dominant effect types. Workers can be submitted as tasks to schedulers like SLURM, LSF, or Google Cloud Batch. This means that the wall (real-world) time increases at a much slower rate than CPU time. Splitting target genomes by chromosome is optional but offers significant performance improvements. For example, splitting UKB data by chromosome means that each worker processes 4.2 million target variants on average, versus 93 million target variants in the combined target genomes.  Splitting by chromosome also enables horizontal scaling (i.e., adding additional workers), which is typically more desirable than vertical scaling (i.e., allocating more RAM / CPU to a worker).

To test the scalability of calculations using pgsc_calc we applied the Calculator to two datasets: 929 samples (78,097,677 variants) from the HGDP[12], and 487,396 samples (93,095,623 variants) from UK Biobank (UKB) imputed using the HRC reference panel.[13] To evaluate the influence of ancestry adjustment on computational resources the the `--run_ancestry` command was run for HGDP using the 1kGP reference panel, and for UKB the HGDP+1kGP reference panel was used. These benchmarks run on multiple sets of scoring files from the May 8, 2024 PGS Catalog data release corresponding to metaGRS$_{CAD}$ (PGS000018) and body mass index (EFO_0004340; 102 scores). To illustrate the effects of PGS adjustment we plot the distributions of PGS000018 (metaGRS$_{CAD}$) in the datasets, after grouping the data on the Most Similar Population assignments.

The impact of genetic ancestry estimation and normalization on performance is largest for small jobs. Most of this work only needs to be done once for each set of target genomes. Recently we have made significant caching improvements to further improve performance and make PGS calculation faster and more environmentally sustainable when calculating new scores on previously processed samples.[14] Work which is stable across multiple calculations, such as preparing target genomes or estimating genetic ancestry, is skipped to save resources (see **Supplementary Figure 6** for marginal resource usage**)**. Previously cached work which has changed - such as applying new scoring files to target genomes - is automatically invalidated by Nextflow without any user input, which causes new processes to be launched.

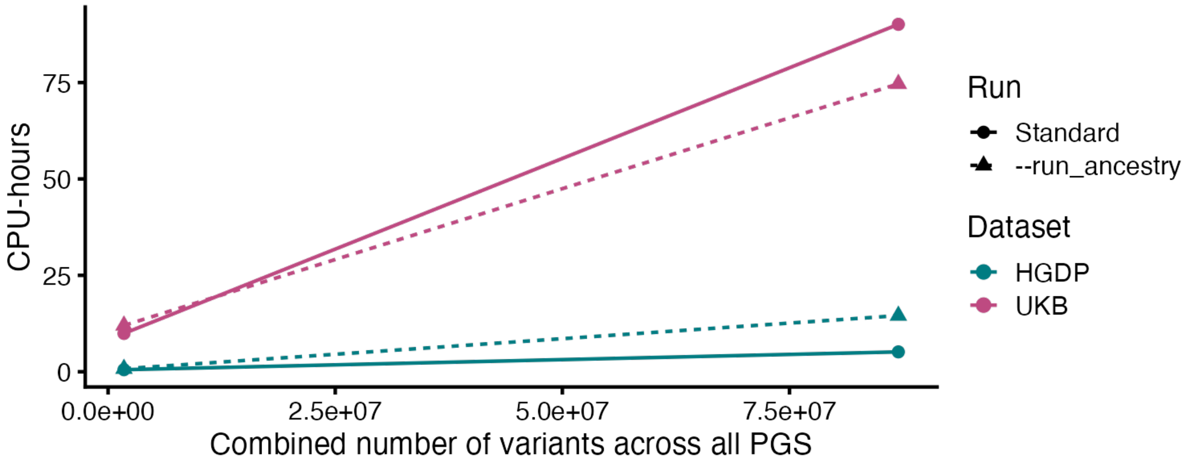


**Supplementary Figure 6. The number of variants in PGS is the main factor that influences scaling.** CPU-time of PGS calculation of PGS000018 and body mass index (EFO_0004340; 102 scores) for 929 HGDP or 487,396 UKB individuals. CPU benchmarks exclude run times for initial data harmonization and ancestry calculation steps and display only the marginal CPU time for new runs of pgsc_calc on the same samples. CPU benchmarking was run on a shared cluster using Intel Xeon Scalable Processors (Platinum 8368Q).

#### Comparison of `pgsc_calc` with other tools

We provide a summary of the key features of the PGS Catalog calculator (`pgsc_calc`) in comparison to other tools (plink2[2], pgs-calc[15], PRScalc[16]) that are are capable of PGS calculation (**Supplementary Table 2**). In contrast to other tools such as pgs-calc[15] and PRScalc[16], the PGS Catalog Calculator is the only PGS calculation tool wholly implemented within a workflow manager and provides additional features to normalize PGS using genetic ancestry. Another pipeline describing the implementation of scoring, PCA, and ancestry adjustment has also been described (eMERGE GIRA; https://github.com/broadinstitute/palantir-workflows/blob/v0.10/ImputationPipeline/ScoringTasks.wdl)[11] in WDL; however, there is little documentation limiting its usability by external users. Our `pgsc_calc` tool also adopts the nf-core community framework to implement best practice such as continuous integration tests, code guidelines, and code templates.[17] Our main justification for using a workflow manager is portability: we aim to make sure that it's possible to bring our code to the users data, because access to human genetic data is strictly controlled and often cannot leave specific storage or compute environments. Workflow managers also bring other benefits: they make it simple to share our versioned code, and help to ensure calculated results reliably reproduce regardless of the underlying compute environment.[18] In contrast, pgs-calc[15] is embedded as a process inside an imputation server, so data must be transferred to a central location before results can be calculated. PRScalc offers a privacy-preserving approach by keeping data client side and calculating PGS using local system resources; however, this approach is less compatible with offline TREs and would not scale to biobank size data.

A limitation of the calculator is that it only implements a command-line interface compared with the graphical user interfaces provided by pgs-calc[15] (via the Michigan Imputation Server[11] website) and PRScalc[16]. However, our schema provides native support for the Seqera Platform (formerly Nextflow Tower) - users that prefer a web interface are able to launch the workflow using the Seqera Platform by linking their HPC or cloud credentials to the platform. Additionally, we provide extensive user-friendly documentation including a getting started tutorial, how-to guides, explanations of PGS, and API reference materials (https://pgsc-calc.readthedocs.io/, https://pygscatalog.readthedocs.io/). An active community discussion forum is also available for users to ask questions about PGS calculation or to report problems they experience (https://github.com/PGScatalog/pgsc_calc/discussions).

It's possible to calculate PGS using other methods, such as using plink2[2] directly, or software libraries like LDpred2.[19] However, these approaches don't provide an end to end solution to PGS calculation and require significant manual work or specialist domain knowledge to operate effectively (e.g. querying the PGS Catalog API, data harmonization). In contrast, the PGS Catalog Calculator requires minimal user input to calculate PGS reliably, and will provide users with clear errors when problems occur during PGS calculation, such as an insufficient match rate between scoring file variants and target genomes.

**Supplementary Table 2. Comparison of `pgsc_calc` to other PGS calculation tools.**

| **Feature** | **PGS Catalog Calculator** (`pgsc_calc`) | `plink2 --score`[2] | `pgs-calc`[15] | `PRScalc`[16] |
|---|---|---|---|---|
| **Target genome input format** | | | | |
| VCF | ✅ | ✅ | ✅ | ❌ |
| PLINK1 binary file format (bed / bim / fam) | ✅ | ✅ | ❌ | ❌ |
| PLINK2 binary file format (pgen / pvar / psam) | ✅ | ✅ | ❌ | ❌ |
| 23andMe format | ❌ | ✅ | ❌ | ✅ |
| **Polygenic score models** | | | | |

| **Feature** | **PGS Catalog Calculator** (`pgsc_calc`) | `plink2 --score`[2] | `pgs-calc`[15] | `PRScalc`[16] |
|---|---|---|---|---|
| Integration with PGS Catalog API to select scores by trait, publication, or PGS ID (in correct genome build) | ✅ | ❌ | | ✅ |
| Support PGS Catalog scoring file format v2 to report score metadata including citations and license | ✅ | ❌ | ✅ | ❌ |
| Automatic variant matching between polygenic score model and target genomes to correctly identify effect alleles despite allele flips, ambiguous alleles, and strand problems | ✅ | ❌ | ✅ | ❌ |
| Auditable log of matched variants and minimum overlap thresholds to minimize user error and misapplication of models | ✅ | ❌ | ✅ | ❌ |
| Automatic liftover of custom polygenic score models between GRCh37 and GRCh38 | ✅ | ❌ | *Not within the tool* | ❌ |
| Custom polygenic score model input | ✅ | ✅ | ❌ | ❌ |
| **Polygenic score calculation** | | | | |
| Automatically combine multiple models to support parallel calculation | ✅ | ❌ | ✅ | ❌ |
| Imputed allele dosages for missing genotypes from reference data | ✅ | ❌ | ❌ | ❌ |
| Polygenic score calculation | ✅ | ✅ | ✅ | ✅ |
| Automatic chromosome parallelization and score aggregation | ✅ | ❌ | *Within pipeline* | ❌ |
| **Genetic ancestry** | | | | |
| Predicted most similar population with robust PCA using a reference panel | ✅ | ❌ | ✅ | ❌ |
| Custom reference panel support | ✅ | ❌ | ❌ | ❌ |
| Adjustment of PGS using genetic ancestry information | ✅ | ❌ | ❌ | ❌ |
| **Outputs & Interface** | | | | |
| Format | text-file | text-file | text-file | json |
| Interface | Command line | Command line | Primarily web-based server (https://imputationserver.sph.umich.edu/), command line tool also available | Web |

| **Feature** | **PGS Catalog Calculator** (`pgsc_calc`) | `plink2 --score`[2] | `pgs-calc`[15] | `PRScalc`[16] |
|---|---|---|---|---|
| User-friendly documentation | Extensive for both command line interface, results, and interpretation: https://pgsc-calc.readthedocs.io/en/latest/index.html | Limited: https://www.cog-genomics.org/plink/2.0/score | Only for web interface: https://imputationserver.readthedocs.io/en/latest/pgs/getting-started/ | Limited, only for interface: https://episphere.github.io/prs/ |
| **Portability** | | | | |
| Natively deploy with Docker containers | ✅ | ❌* | ❌ | **NA** |
| Natively deploy with Singularity containers | ✅ | ❌* | ❌ | **NA** |
| Natively deploy with Anaconda environments | ✅ | ❌* | ❌ | **NA** |
| Native support for HPC deployment (e.g. SLURM, LSF) | ✅ | ❌ | ❌ | **NA** |
| Native support for cloud deployment (e.g. Google Cloud Batch) | ✅ | ❌ | ❌ | **NA** |
| Supports offline / airlocked environments (e.g. Trusted Research Environments) | ✅ | ✅ | ✅ | ❌ |

*available via the biocontainers[20] project
